# Clinical characteristics and mortality associated with COVID-19 in Jakarta, Indonesia: a hospital-based retrospective cohort study

**DOI:** 10.1101/2020.11.25.20235366

**Authors:** Henry Surendra, Iqbal RF Elyazar, Bimandra A Djaafara, Lenny L Ekawati, Kartika Saraswati, Verry Adrian, Widyastuti, Dwi Oktavia, Ngabila Salama, Rosa N Lina, Adhi Andrianto, Karina D Lestari, Erlina Burhan, Anuraj H Shankar, Guy Thwaites, J. Kevin Baird, Raph L. Hamers

## Abstract

**Background:** Data on COVID-19-related mortality and associated factors from low-resource settings are scarce. This study examined clinical characteristics and factors associated with in-hospital mortality of COVID-19 patients in Jakarta, Indonesia, from March 2 to July 31, 2020.

**Methods:** This retrospective cohort included all hospitalised patients with PCR-confirmed COVID-19 in 55 hospitals. We extracted demographic and clinical data, including hospital outcomes (discharge or death). We used Cox regression to examine factors associated with mortality.

**Findings:** Of 4265 patients with a definitive outcome by July 31, 3768 (88%) were discharged and 497 (12%) died. The median age was 46 years (IQR 32–57), 5% were children, and 31% had at least one comorbidity. Age-specific mortalities were 11% (7/61) for <5 years; 4% (1/23) for 5-9; 2% (3/133) for 10-19; 2% (8/638) for 20-29; 3% (26/755) for 30-39; 7% (61/819) for 40-49; 17% (155/941) for 50-59; 22% (132/611) for 60-69; and 34% (96/284) for ≥70. Risk of death was associated with higher age; pre-existing hypertension, cardiac disease, chronic kidney disease or liver disease; clinical diagnosis of pneumonia; multiple (>3) symptoms; and shorter time from symptom onset to admission. Patients <50 years with >1 comorbidity had a nearly six-fold higher risk of death than those without (adjusted hazard ratio 5·50, 95% CI 2·72-11·13; 27% vs 3% mortality).

**Interpretation:** Overall mortality was lower than reported in high-income countries, probably due to younger age distribution and fewer comorbidities. However, deaths occurred across all ages, with >10% mortality among children <5 years and adults >50 years.

## Background

Severe acute respiratory syndrome coronavirus 2 (SARS-CoV-2), the virus that causes coronavirus disease 2019 (COVID-19), has spread rapidly around the world since it was first reported in Wuhan, China in December 2019 [1]. Current understanding of COVID-19 mortality mostly comes from clinical epidemiological studies conducted in the early phase of the pandemic in China [2–4], and in high-income countries of North America [5–8] and Europe [9,10]. Severe outcomes of COVID-19 in those settings have been consistently associated with older age and pre-existing chronic conditions, such as hypertension, diabetes, obesity, cardiac disease, chronic kidney disease, and liver disease [2,4–10].

However, most cases to date have occurred in low-and middle-income countries (LMIC) [11], where differences in age distribution, comorbidities, access to quality health services, and other factors, may greatly influence trends regarding severe outcomes, but data are limited [12–17]. Indonesia is the fourth most populous country (population 274 million) and the LMIC that has suffered the highest number of COVID-19 confirmed cases and deaths in Southeast Asia, second only to India in Asia [11]. Since the first two laboratory-confirmed SARS-CoV-2 infections were reported on March 2, 2020, Indonesia has reported a total of 478 720 cases and 15 503 deaths (3·2% confirmed case fatality rate) up to November 18, 2020 [18], with the highest numbers of reported cases (25%, 121 818) and deaths (16%, 2470) in the capital city of Jakarta. COVID-19 cases and deaths in Jakarta rapidly escalated during the first two months of the outbreak (March-April 2020), and have steadily trended upward through November 2020 [18].

Indonesia has high burdens of major infectious diseases like malaria, tuberculosis, HIV and other tropical infections [19], as well as non-communicable diseases, with an estimated 73% of deaths caused predominantly by cardiovascular diseases, cancers, chronic pulmonary diseases, diabetes, and others [20]. As in many LMIC, substantial proportions of the population face barriers in accessing quality health care services due to under-resourced and fragile health systems [12], often leaving underlying chronic comorbidities unrecognised and/or poorly managed [21]. These factors may aggravate chronic non-communicable diseases and worsen COVID-19 patient outcomes.

Studies of COVID-19-related mortality in Asia [16,22,23], and in particular from low-resource settings have been limited. Given that several major urban centres of Southeast Asia like Bangkok, Phnom Penh, Ho Chi Minh City, and Kuala Lumpur, have thus far been spared major COVID-19 epidemics, the explosive epidemic in Jakarta can provide insights directly relevant to similar settings in other LMIC. To this end, we analysed the complete clinical epidemiological surveillance data from the Jakarta Health Office, reporting on admissions to 55 COVID-19-designated hospitals within the city, during the first five months of the epidemic (March through July 2020).

## Methods

### Study design and participants

This was a retrospective cohort study to assess demographic and clinical factors associated with mortality of adults and children hospitalised with COVID-19 in Jakarta, Indonesia. The study population included all PCR-confirmed COVID-19 patients recorded by the Jakarta Health Office who either died or were discharged alive between March 2 and July 31, 2020. In accordance with Indonesia’s national COVID-19 guidelines [24], confirmatory SARS-CoV-2 PCR testing was conducted on mouth and/or throat swab specimens in COVID-19 reference laboratories, and patients were discharged from hospital after two consecutive PCR-negative tests.

### Data collection

The study data were extracted from the Jakarta Health Office official COVID-19 epidemiological investigation form. In-hospital mortality was duly reported by the surveillance officer at each hospital. Data regarding dates of onset of illness, SARS-CoV-2 PCR testing, hospital admission, and outcomes (discharge or death) were recorded, along with age, sex, symptoms, comorbidities, and some critical indicators (e.g. immediate admission to intensive care unit). Comorbidities were recorded at admission by attending clinical staff, either based on clinical assessments or patient reporting. Fever was defined as axillary temperature of at least 38°C [24]. Clinical diagnosis of pneumonia (indicative of lower respiratory tract involvement) was established by the treating physician, based on clinical and radiological evaluation [24].

### Statistical analysis

Descriptive statistics included proportions for categorical variables and medians and interquartile ranges (IQRs) for continuous variables. We calculated time in days from symptom onset to hospital admission, and length of hospital stay until death or discharge. We used the Mann-Whitney U test, χ^2^ test, or Fisher’s exact test to compare characteristics between deceased and discharged patients. Graphs of Nelson-Aalen cumulative hazard function were done to compare time-dependent mortality risks based on demographic and clinical risk factors. We used univariable and multivariable Cox regression models to determine the risk of death, expressed as hazard ratio with 95% confidential intervals. All independent variables with p-value <0.10 in univariable analysis were included in the multivariable models. Final model selection was informed by likelihood ratio tests. No imputations were made for missing data, and because of the high proportion of missing data for obesity (53%) this variable was not included in regression analysis. We reported two final multivariable Cox models. In the first model, we assessed the effect of each type of comorbidity (hypertension, diabetes, cardiac disease, chronic kidney disease and liver disease), and, in the second model, the number of comorbidities (0, 1 or >1), along with other demographic and clinical factors. We used interaction terms to examine potential effect modification by age and sex. We confirmed the statistical assumptions were met for proportional hazards analysis. We set statistical significance at 0·05, and all tests were two sided. All analyses were done in Stata/IC 15.1 (StataCorp, College Station, TX, USA). This study is reported as per Strengthening the Reporting of Observational Studies in Epidemiology (STROBE) guidelines [25].

### Role of the funding source

The funder of the study had no role in study design, data collection, data analysis, data interpretation, or writing of the report. The corresponding author had full access to all of the data and the final responsibility to submit for publication.

### Ethics

This study was approved by the Health Research Ethics Committee of the National Institute of Health Research and Development, Ministry of Health Indonesia (LB.02.02/2/KE.554/2020). The requirement for patient consent was waived as this was a secondary analysis of anonymised routine surveillance data.

## Results

Between March 2 and July 31, 2020, a total of 21 397 PCR-confirmed COVID-19 cases were recorded by the Jakarta Health Office (Figure 1). Of those, 4522 (21%) were admitted to hospital because of symptomatic disease and 4265 (94%) had reached a definitive outcome no later than July 31, 2020, i.e. deceased or discharged, and were therefore included in this analysis. There were 257 (6%) who were still hospitalised. The study flow chart and completeness of key data are presented in Figure 1.

**Figure 1:**
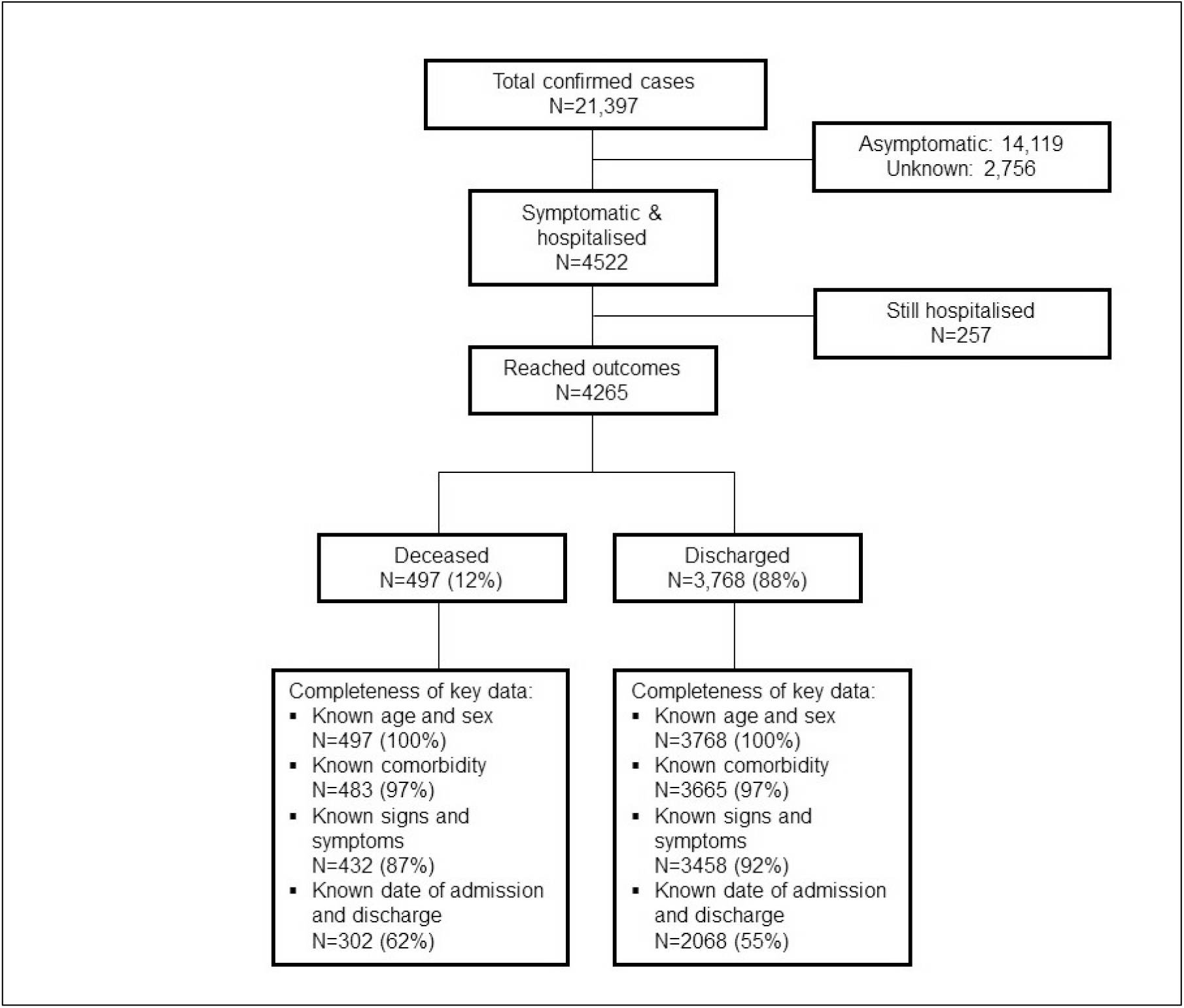
Study flow chart and completeness of key variables.

Table 1 presents the characteristics of the 4265 patients included in the analysis. The median age was 46 years (IQR 32-57, range 0·1-99), 217 (5%) were children, and 2217 (52%) were men. The most common presenting symptom was cough (66%, 2788), followed by fever (53%, 2192), malaise (35%, 1460), shortness of breath (32%, 1335), and others (Figure 2A). The median number of symptoms was 3 (IQR 2-5, range 1-12), 1458 (39%) patients had more than three symptoms. We found a moderate degree of overlap between the three most common symptoms (Figure 2C). Overall, 31% (1299) of patients had a record of one or more pre-existing comorbidities, including hypertension (19%, 795), diabetes mellitus (12%, 501), cardiac disease (10%, 392), chronic obstructive pulmonary disease (4%, 178), chronic kidney disease (3%, 108), and others (Figure 2B). There was little overlap between the three most common comorbidities (Figure 2D). There were 1488 (37%) patients with a clinical diagnosis of pneumonia. Among 3214 patients with complete records, 102 (3%) patients were directly admitted to ICU, and among 3211 patients with complete records, 55 (2%) were intubated for mechanical ventilation upon admission.

**Table 1:**
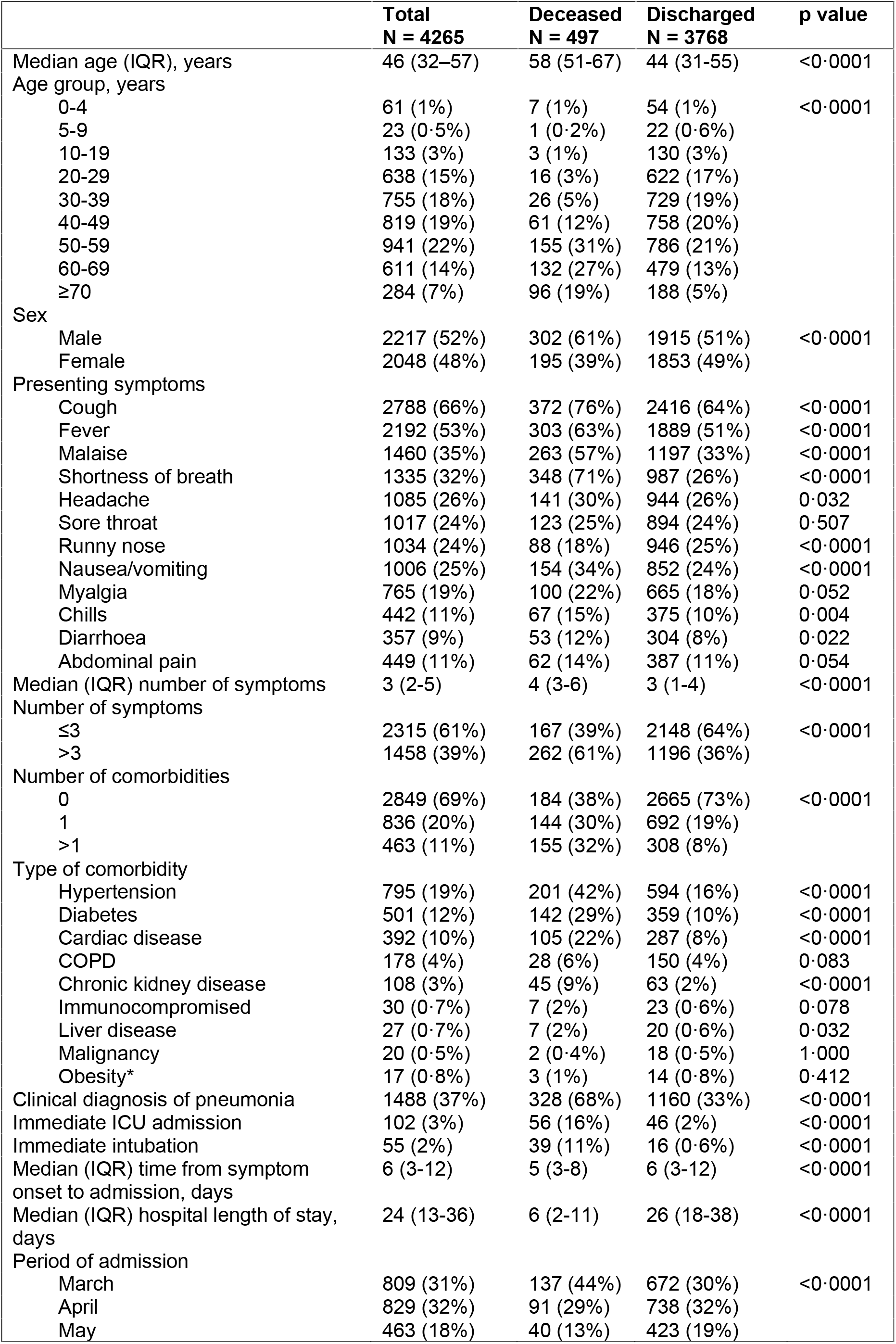
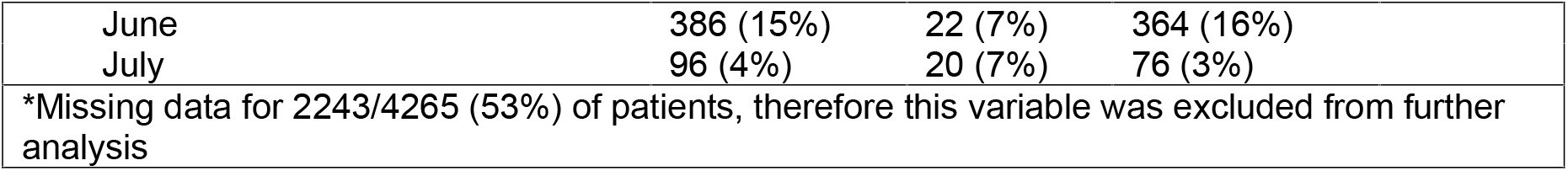
Demographics, clinical characteristics and outcomes of COVID-19 hospitalised patients in Jakarta, Indonesia.

|  | **Total**<br>**N = 4265** | **Deceased**<br>**N = 497** | **Discharged**<br>**N = 3768** | **p value** |
| --- | --- | --- | --- | --- |
| Median age (IQR), years | 46 (32–57) | 58 (51–67) | 44 (31–55) | <0.0001 |
| Age group, years |  |  |  |  |
| 0–4 | 61 (1%) | 7 (1%) | 54 (1%) | <0.0001 |
| 5–9 | 23 (0.5%) | 1 (0.2%) | 22 (0.6%) |  |
| 10–19 | 133 (3%) | 3 (1%) | 130 (3%) |  |
| 20–29 | 638 (15%) | 16 (3%) | 622 (17%) |  |
| 30–39 | 755 (18%) | 26 (5%) | 729 (19%) |  |
| 40–49 | 819 (19%) | 61 (12%) | 758 (20%) |  |
| 50–59 | 941 (22%) | 155 (31%) | 786 (21%) |  |
| 60–69 | 611 (14%) | 132 (27%) | 479 (13%) |  |
| ≥70 | 284 (7%) | 96 (19%) | 188 (5%) |  |
| Sex |  |  |  |  |
| Male | 2217 (52%) | 302 (61%) | 1915 (51%) | <0.0001 |
| Female | 2048 (48%) | 195 (39%) | 1853 (49%) |  |
| Presenting symptoms |  |  |  |  |
| Cough | 2788 (66%) | 372 (76%) | 2416 (64%) | <0.0001 |
| Fever | 2192 (53%) | 303 (63%) | 1889 (51%) | <0.0001 |
| Malaise | 1460 (35%) | 263 (57%) | 1197 (33%) | <0.0001 |
| Shortness of breath | 1335 (32%) | 348 (71%) | 987 (26%) | <0.0001 |
| Headache | 1085 (26%) | 141 (30%) | 944 (26%) | 0.032 |
| Sore throat | 1017 (24%) | 123 (25%) | 894 (24%) | 0.507 |
| Runny nose | 1034 (24%) | 88 (18%) | 946 (25%) | <0.0001 |
| Nausea/vomiting | 1006 (25%) | 154 (34%) | 852 (24%) | <0.0001 |
| Myalgia | 765 (19%) | 100 (22%) | 665 (18%) | 0.052 |
| Chills | 442 (11%) | 67 (15%) | 375 (10%) | 0.004 |
| Diarrhoea | 357 (9%) | 53 (12%) | 304 (8%) | 0.022 |
| Abdominal pain | 449 (11%) | 62 (14%) | 387 (11%) | 0.054 |
| Median (IQR) number of symptoms | 3 (2–5) | 4 (3–6) | 3 (1–4) | <0.0001 |
| Number of symptoms |  |  |  |  |
| ≤3 | 2315 (61%) | 167 (39%) | 2148 (64%) | <0.0001 |
| >3 | 1458 (39%) | 262 (61%) | 1196 (36%) |  |
| Number of comorbidities |  |  |  |  |
| 0 | 2849 (69%) | 184 (38%) | 2665 (73%) | <0.0001 |
| 1 | 836 (20%) | 144 (30%) | 692 (19%) |  |
| >1 | 463 (11%) | 155 (32%) | 308 (8%) |  |
| Type of comorbidity |  |  |  |  |
| Hypertension | 795 (19%) | 201 (42%) | 594 (16%) | <0.0001 |
| Diabetes | 501 (12%) | 142 (29%) | 359 (10%) | <0.0001 |
| Cardiac disease | 392 (10%) | 105 (22%) | 287 (8%) | <0.0001 |
| COPD | 178 (4%) | 28 (6%) | 150 (4%) | 0.083 |
| Chronic kidney disease | 108 (3%) | 45 (9%) | 63 (2%) | <0.0001 |
| Immunocompromised | 30 (0.7%) | 7 (2%) | 23 (0.6%) | 0.078 |
| Liver disease | 27 (0.7%) | 7 (2%) | 20 (0.6%) | 0.032 |
| Malignancy | 20 (0.5%) | 2 (0.4%) | 18 (0.5%) | 1.000 |
| Obesity* | 17 (0.8%) | 3 (1%) | 14 (0.8%) | 0.412 |
| Clinical diagnosis of pneumonia | 1488 (37%) | 328 (68%) | 1160 (33%) | <0.0001 |
| Immediate ICU admission | 102 (3%) | 56 (16%) | 46 (2%) | <0.0001 |
| Immediate intubation | 55 (2%) | 39 (11%) | 16 (0.6%) | <0.0001 |
| Median (IQR) time from symptom onset to admission, days | 6 (3–12) | 5 (3–8) | 6 (3–12) | <0.0001 |
| Median (IQR) hospital length of stay, days | 24 (13–36) | 6 (2–11) | 26 (18–38) | <0.0001 |
| Period of admission |  |  |  |  |
| March | 809 (31%) | 137 (44%) | 672 (30%) | <0.0001 |
| April | 829 (32%) | 91 (29%) | 738 (32%) |  |
| May | 463 (18%) | 40 (13%) | 423 (19%) |  |

|  |  |  |  |
| --- | --- | --- | --- |
| June | 386 (15%) | 22 (7%) | 364 (16%) |
| July | 96 (4%) | 20 (7%) | 76 (3%) |
| *Missing data for 2243/4265 (53%) of patients, therefore this variable was excluded from further analysis |  |  |  |

**Figure 2:**
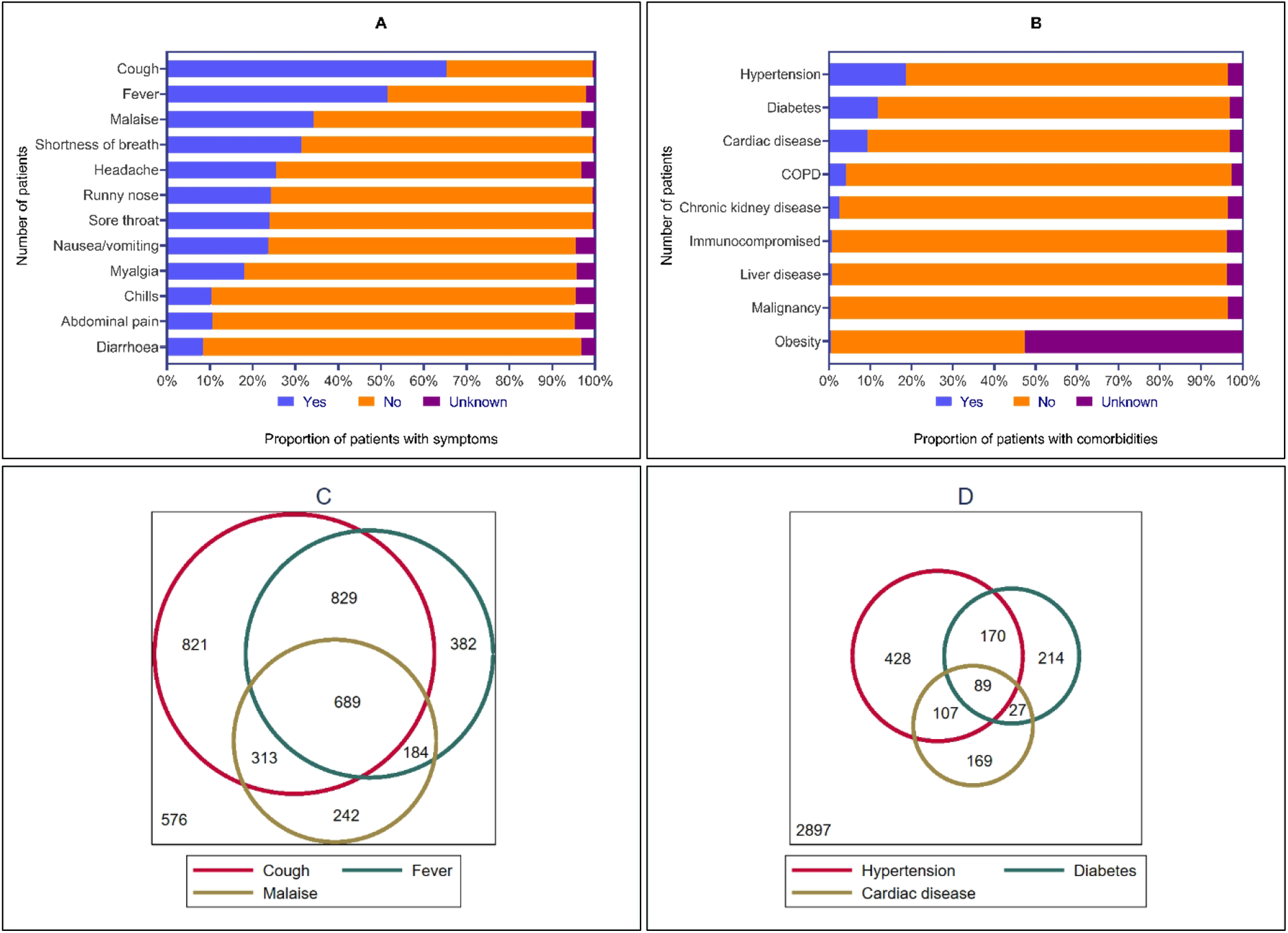
Presenting symptoms and comorbidities in patients hospitalised with COVID-19 in Jakarta. Symptoms (A) and comorbidities (B) by frequency (see Table 1 for values); Scaled Euler diagrams of overlap of commonest symptoms (C) and comorbidities

Of 4265 patients with a known outcome, 497 (12%) had died, and 3768 (88%) were discharged alive. 36 (7%) deceased patients had been declared dead at hospital arrival and were censored from the Cox analysis. The highest numbers of deaths were observed between March 28 and April 4, 2020 (Figure 3A). The median time from symptom onset to admission was 6 days (IQR 3-12) overall, 5 days (IQR 3-8) among deceased patients, and 6 days (IQR 3-12) among those discharged. The median length of hospital stay was 24 days (IQR 13-36) overall, 6 days (IQR 2-11) among deceased patients, and 26 days (IQR 18-38) among survivors.

**Figure 3:**
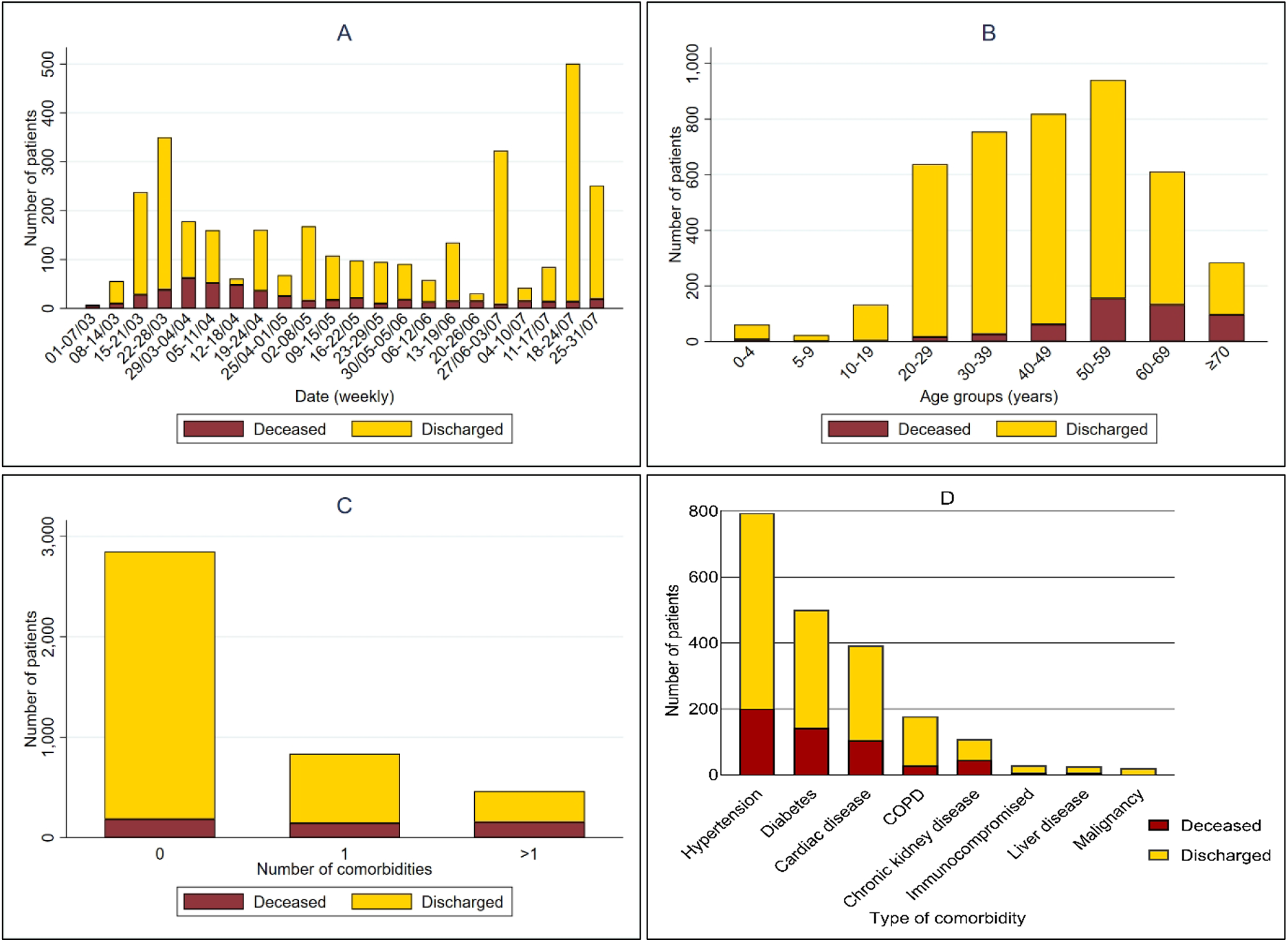
Outcomes of hospitalization over time, by age group and pre-existing comorbidity. Proportions of in-hospital deaths and discharges over time (A), by age groups (B), by number of pre-existing comorbidities (C), and by type of comorbidity (D).

Compared to discharged patients, deceased patients were older (median 58 vs 44 years); more likely to be men (14% vs 10%); more likely to have presenting symptoms of cough, fever, malaise, shortness of breath, headache, nausea/vomiting, chills, diarrhoea, abdominal pain, and >3 symptoms (Table 1); more likely to have a clinical diagnosis of pneumonia (68% vs 33%); to be directly admitted to intensive care (16% vs 2%); to receive mechanical ventilation at admission (11% vs 0·6%); and more likely to have a history of any comorbidity (62% vs 27%), 1 comorbidity (30% vs 19%) or >1 comorbidities (32% vs 8%), specifically hypertension (42% vs 16%), diabetes (29% vs 10%), cardiac disease (22% vs 8%), chronic kidney disease (9% vs 2%), and liver disease (2% vs 0·6%) (Table 1; Figure 3C and 3D).

Although a large majority of deaths (78%, 383) was 50 years or older, death occurred across all age groups. Age-specific mortalities were 11% (7/61) for 0-4 years; 4% (1/23) for 5-9 years; 2% (3/133) for 10-19 years; 3% (8/638) for 20-29 years; 3% (26/755) for 30-39 years; 7% (61/819) for 40-49 years; 16% (155/941) for 50-59 years; 22% (132/611) for 60-69 years; 34% (96/284) for ≥70 years (Supplementary Table 1 and Figure 3B).

Among the seven children below 5 years who died with COVID-19, four (57%) were boys, with an age range of 0 to 3 years; four (57%) had a clinical diagnosis of pneumonia, four (67%) had >3 symptoms at presentation (1 unknown), two (29%) had a known underlying condition (i.e. cardiac disease and immunocompromised status), and two (29%) were directly admitted to ICU for mechanical ventilation, and five (71%) died within a week after admission. Further details on the clinical course, disease management, underlying conditions and the exact cause of death were not available for this analysis.

The mortality increased by age and number of pre-existing comorbidities (Supplementary Figure 1). The mortality for the age group <50 years was 5% (110/2372) overall, 3% (59/1945) for those without any comorbidity, 8% (25/331) for those with 1 comorbidity, and 27% (26/96) for those with >1 comorbidities; for the age group 50-59 years 16% (146/2372), 11% (56/515), 18% (42/231), and 32% (48/151), respectively; for the age group 60-69 years 22% (132/601), 16% (47/286), 25% (46/181), and 29% (39/134), respectively; and for the age group ≥70 years 34% (95/278), 21% (22/103), 33% (31/93), and 51% (42/82), respectively.

The cumulative incidence of death suggested that almost all deaths occurred within 30 days of admission and differed by demographic and clinical factors (Figure 4). In univariable Cox analysis (Supplementary Table 2), the risk of death was associated with older age, sex, number of pre-existing comorbidities, hypertension, diabetes, cardiac disease, chronic kidney disease, liver disease, number of presenting symptoms, pneumonia, immediate ICU admission and intubation, calendar period of admission, and duration from symptom onset to hospital admission.

**Figure 4:**
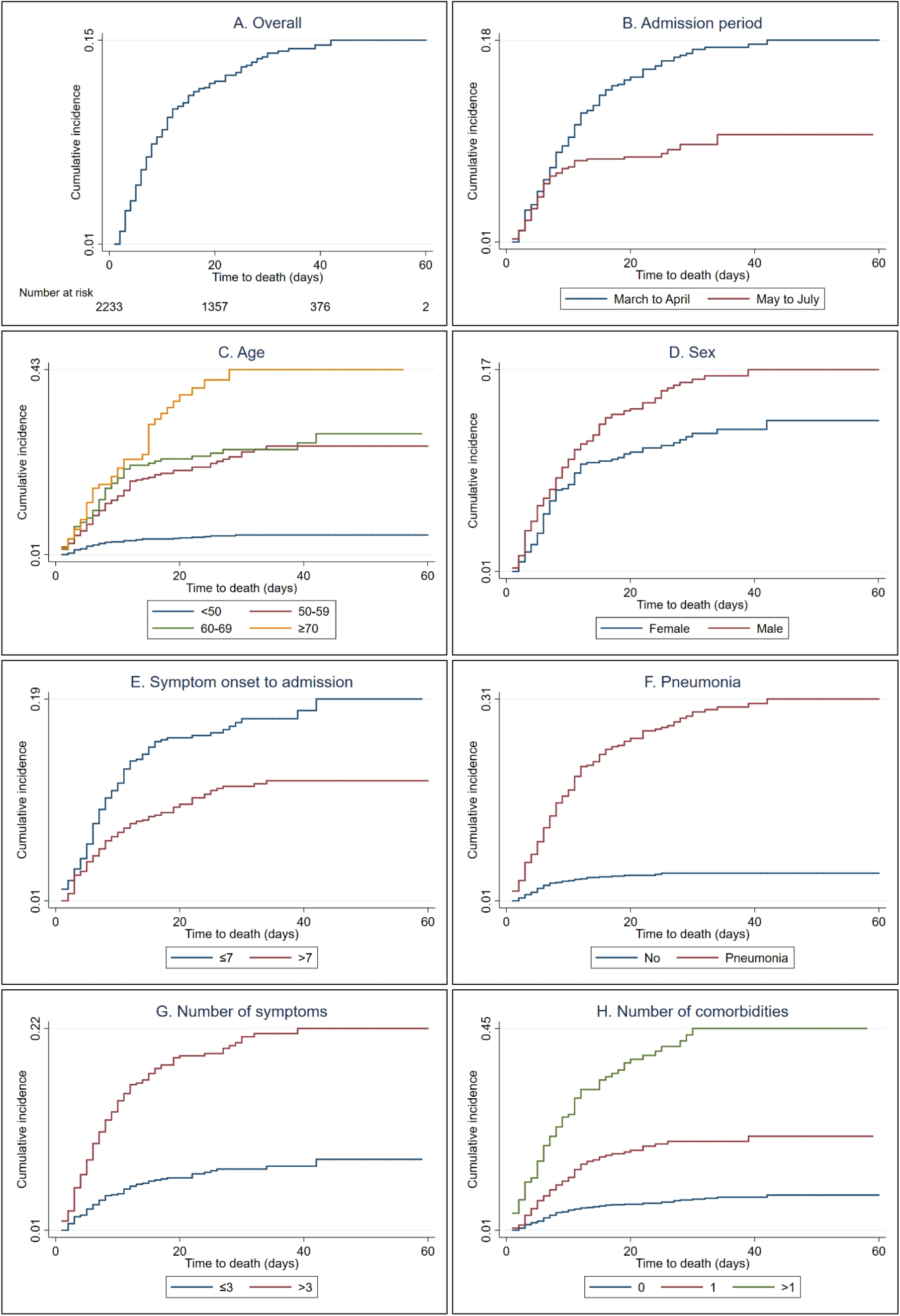
Nelson-Aalen cumulative hazard plots. (A) overall time-dependent risk of death. Comparison of the time-dependent risk of death by period of admission (B), age groups (C), sex (D), duration from symptom onset to hospital admission (E), clinical diagnosed pneumonia (F), number of symptoms (G), and by number of comorbidities (H). The analysis was conducted among patients with known admission and discharge dates, with maximum 60 days of follow up time.

In the first multivariable Cox model (Figure 5A), the risk of death was increased for age groups 50-59 years (aHR 2·47, 95%CI 1·68-3·61), 60-69 years (aHR 2·19, 95%CI 1·45-3·32), and ≥70 years (aHR 3·30, 95%CI 2·06-5·29), compared to <50 years; for patients with pre-existing hypertension (aHR 1·38, 95%CI 1·01-1·88), cardiac disease (aHR 1·58, 95%CI 1·09-2·31), chronic kidney disease (aHR 1·69, 95%CI 1·01-2·83), or liver disease (aHR 4·02, 95%CI 1·15-14·03); for patients who had pneumonia (aHR 3·91, 95%CI 2·68-5·69), >3 symptoms at presentation (aHR 1·60, 95%CI 1·16-2·21; reference ≤3 symptoms), or duration of ≤7 days from symptom onset to hospital admission (aHR 1·57, 95%CI 1·18-2·09; reference >7 days).

**Figure 5:**
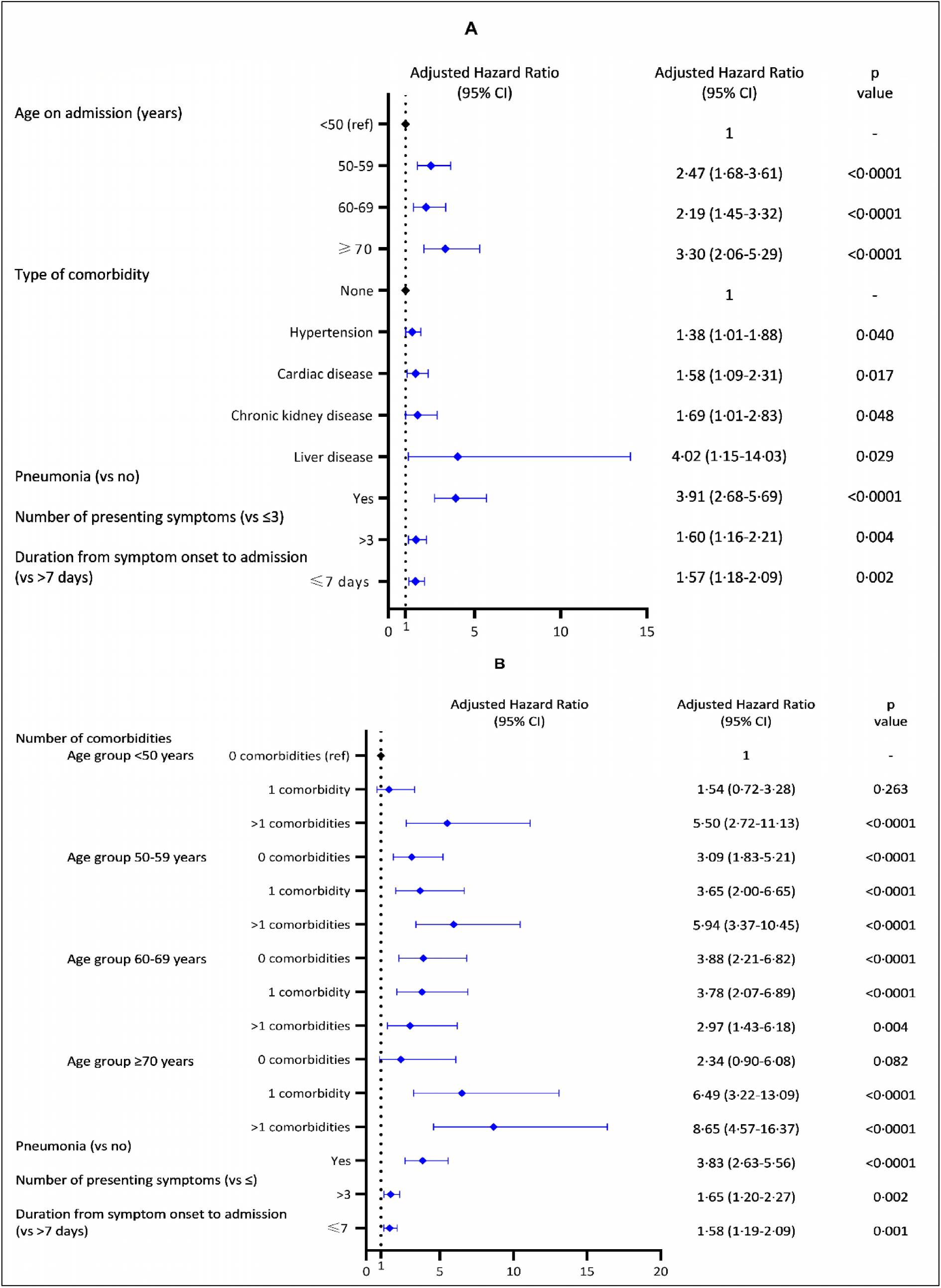
Multivariable Cox proportional hazard models. The models present the demographic and clinical risk factors independently associated with in-hospital death (A), and adjusted age-specific effect of number of comorbidities on in-hospital death (B). Thirty-six patients who had died on hospital arrival were censored from the analysis. Both models did not violate the proportional-hazards assumption (p=0·089 for model 1 and p=0·084 for model 2). Sex, date of admission and diabetes were assessed in the multivariable models but were not associated with in-hospital death.

In the second multivariable Cox model (Figure 5B), we found that the effect of comorbidities on death risk was modified by age. Compared to age group <50 years without comorbidity, the risk of death was increased for age <50 years with >1 comorbidities (aHR 5·50, 95% CI 2·72-11·13); for 50-59 years without comorbidity (aHR 3·09, 95%CI 1·83-5·21), with 1 comorbidity (aHR 3·65, 95%CI 2·00-6·65), or with >1 comorbidities (aHR 5·94, 95%CI 3·37-10·45); for 60-69 years without comorbidity (aHR 3·88, 95%CI 2·21-6·82), with 1 comorbidity (aHR 3·78, 95%CI 2·07-6·89), with >1 comorbidities (aHR 2·97, 95%CI 1·43-6·18); for ≥70 years with 1 comorbidity (aHR 6·49, 95%CI 3·22-13·09) or >1 comorbidities (aHR 8·65, 95%CI 4·57-16·37). Both multivariable models found no associations for sex, diabetes, and calendar period of hospital admission. Sex was not found to be an effect modifier.

## Discussion

This retrospective hospital-based study described the complete epidemiological surveillance data of the Jakarta Health Office, including 4265 adults and children with confirmed COVID-19 admitted in 55 COVID-19-designated referral hospitals, during the first five months of the SARS-CoV-2 epidemic. This analysis represents the largest patient series hospitalised with COVID-19 in Southeast Asia, and one of the largest from LMIC to date. The observed disease pattern broadly reflects that reported globally, with patients usually presenting with multiple symptoms of fever, cough, malaise and/or shortness of breath, and nearly 40% having a clinical diagnosis of pneumonia at admission. Overall mortality was 12% (497/4265), and deaths occurred across all ages. Although the majority (57%) of all admissions were younger than 50 years, including 5% children, the large majority (78%) of people who died were 50 years or older. Mortality increased with age, from 7% in patients aged 40-49 years to 34% in patients aged ≥70 years. Mortality among children below 5 years was unexpectedly high at 11% (7/61), which has not been reported elsewhere, although available information on any underlying conditions and the exact cause of death of those children was incomplete. Elevated risk of in-hospital death was independently associated with higher age, history of hypertension, cardiac disease, chronic kidney disease, or liver disease, pneumonia, multiple (>3) symptoms at presentation, and shorter (≤7) duration between symptom onset and hospital admission. The increased risk of in-hospital death associated with advancing age was further augmented by the presence of one or more chronic comorbidities, which were recorded to be present in 31% of patients.

The overall COVID-19-related mortality in Jakarta (12%) was substantially lower than those reported in high-income countries, for example, in the US (21%) [7] and the UK (26%) [9], but those populations were substantially older, with more comorbidities and more frequent presentation with severe disease. By contrast, a nationwide analysis of patients hospitalised with COVID-19 in China reported an overall mortality of just 3·1%, but in that population, severe cases and comorbidities were relatively infrequent, while the median age was similar to Jakarta [4]. Compared with other settings, we found age-specific mortality rates in Jakarta to be similar for patients <50 years (5% in Jakarta, 5% in the US [7], and 4% in the UK [9]), but slightly lower for patients ≥50 years (21% in Jakarta, 27% in the US [7], and 29% in the UK [9]. Given that older age has been consistently associated with severe disease and mortality in patients with COVID-19 [2,5–7,9], the lower median age (46 years) in Jakarta compared to studies from North America and Europe probably accounts for most of the differences observed in overall in-hospital mortality. The younger age at admission is likely to be mainly related to a younger age distribution in the general population in Jakarta, compared to Europe, the US, and China. Nonetheless, we do not dismiss distinct behavioural factors related to risk of infection, for example adherence to preventive measures, mobility, and health seeking, as also reported in India [16].

National surveillance data in Indonesia to date reported a case fatality rate in children aged 0-5 years of 1.0% (123/11 916) [18]. This is much higher than a case fatality rate of 0·16% in children below 5 years of age in a recent large epidemiological study in Southern India [16]. Several other studies in China [26], Brazil [13,27], Uganda [14], and South Africa [15] have reported COVID-19-related deaths among children under 5 years to be rare. A review of data from US, Korea and Europe early in the epidemic estimated the overall case fatality rate among children (0-19 years old) to be low at 0·03 per 100 000 (44/42 846) [28]. Although children appear generally less severely affected than older individuals [29], COVID-19 in children can also be characterized by rapid progressive deterioration leading to death, especially in children with comorbidities. Notably, increasing reports from both high-income and LMIC have described a multisystem inflammatory syndrome in children (MIS-C) associated with COVID-19, which can lead to serious illness [30]; to date, to our knowledge, no such cases have been reported in Indonesia. Factors that explain the high mortality observed among children in Jakarta hospitals may be various, including treatment delays, limited paediatric intensive care capacity, presence of underlying conditions, such as malignancy or malnutrition, among other factors. Further investigations are planned to confirm the extent and cause of death among young children with COVID-19 in Jakarta.

Evidence from previous studies suggests that underlying comorbidities, including pre-existing cardiovascular or cerebrovascular diseases, hypertension, and diabetes mellitus, were associated with poorer COVID-19 outcomes [4,7–9]. Chronic comorbidities among COVID-19 patients were less frequent in Jakarta (31% of patients) than reported in large patient series in North America [7,8], South America [13], and Europe [9]. This could reflect the relatively young population admitted to Jakarta hospitals, but also under-reporting of comorbidities by patients, or under-diagnosis due to variable access to quality health services. Presence of one or more comorbidities, especially pre-existing hypertension, cardiac disease, chronic kidney disease or liver disease strongly increased the risk of death in Jakarta, by 40%, 60%, 70% and 300%, respectively, compared to those without such comorbidities. These findings were consistent with similar studies elsewhere [4,8,9]. In contrast, similar to a study in the US [8], we found no association between diabetes and mortality in Jakarta, whereas studies in China [4] and the UK [9] did so. Obesity has also been recognised as an important risk factor of mortality [9,17], but we could not assess that for lack of complete data.

In Jakarta, the median time from symptom onset to death (median 11 days) was shorter than reported in hospital-based studies from Japan (17 days) [22], China (18·5 days) [2], and the US (12·7 days) [6]; the relatively short in-hospital trajectory towards clinical deterioration and death (median 6 days) could suggest possible delays in hospital admission and treatment. Among survivors, the length of hospital stay was much longer (median 26 days) than reported in other studies [2,6,7,9,22], which is most likely due to the requirement of two consecutive PCR-negative test results prior to discharge, with results delayed due to limited PCR testing capacity in Jakarta during the early phase of the epidemic.

This study had some limitations. The retrospective design and reliance on routine hospital surveillance data meant that, for some key baseline variables, data were incomplete or uniformly unavailable (e.g. vital signs, TB and HIV co-infection, radiology examination, routine laboratory results). Comorbidities were often self-reported or could be under-diagnosed, potentially resulting in underreporting and hence underestimation of effect sizes. Details on supportive care and treatment received, particularly respiratory support and management of secondary infections, were also not available for this analysis. Findings from hospitals reported here may not reflect the mortality rate and risk factors associated with COVID-19-related mortality in the general population.

In conclusion, risk factors associated with in-hospital mortality in Jakarta, Indonesia are broadly similar to those in more developed settings in North America, Europe, and Asia, dominated by advanced age and comorbidities. Lower overall mortality in Jakarta was likely driven by the lower median age of the hospital population. While children represented just 5% of admissions, the 11% mortality that occurred among children below 5 years should prompt further investigation of them as potentially highly vulnerable in LMIC settings. The findings highlight the need for enhanced context-specific public health action to reduce infection risk among vulnerable populations and improved care and treatment for the diseased.

## Data Availability

After publication, the datasets used for this study will be made available to others on reasonable requests to the corresponding author, including a detailed research proposal, study objectives and statistical analysis plan. Deidentified participant data will be provided after written approval from the corresponding author and the Jakarta Health Office.

## Contributors

HS designed the study, cleaned the data, did the analysis, and had full access to all of the data in the study and take responsibility for the integrity of the data and the accuracy of the data analysis. VA, W, DO, and NS collected and verified the data. HS, IRFE, JKB, and RLH drafted the paper. BAD, LLE, KS, VA, W, DO, NS, AA, RNL, KDL, EB, AHS, and GT critically revised the manuscript for important intellectual content and all authors gave final approval for the version to be published.

## Conflict of interests

We declare no competing interests.

## Data sharing statement

### Acknowledgments

This work was funded by the Wellcome Trust, UK (106680/Z/14/Z). We acknowledge all health care workers involved in the diagnosis and treatment of patients, as well as those involved in the field data collection and entry.

## Supplementary data

**Figure S1:**
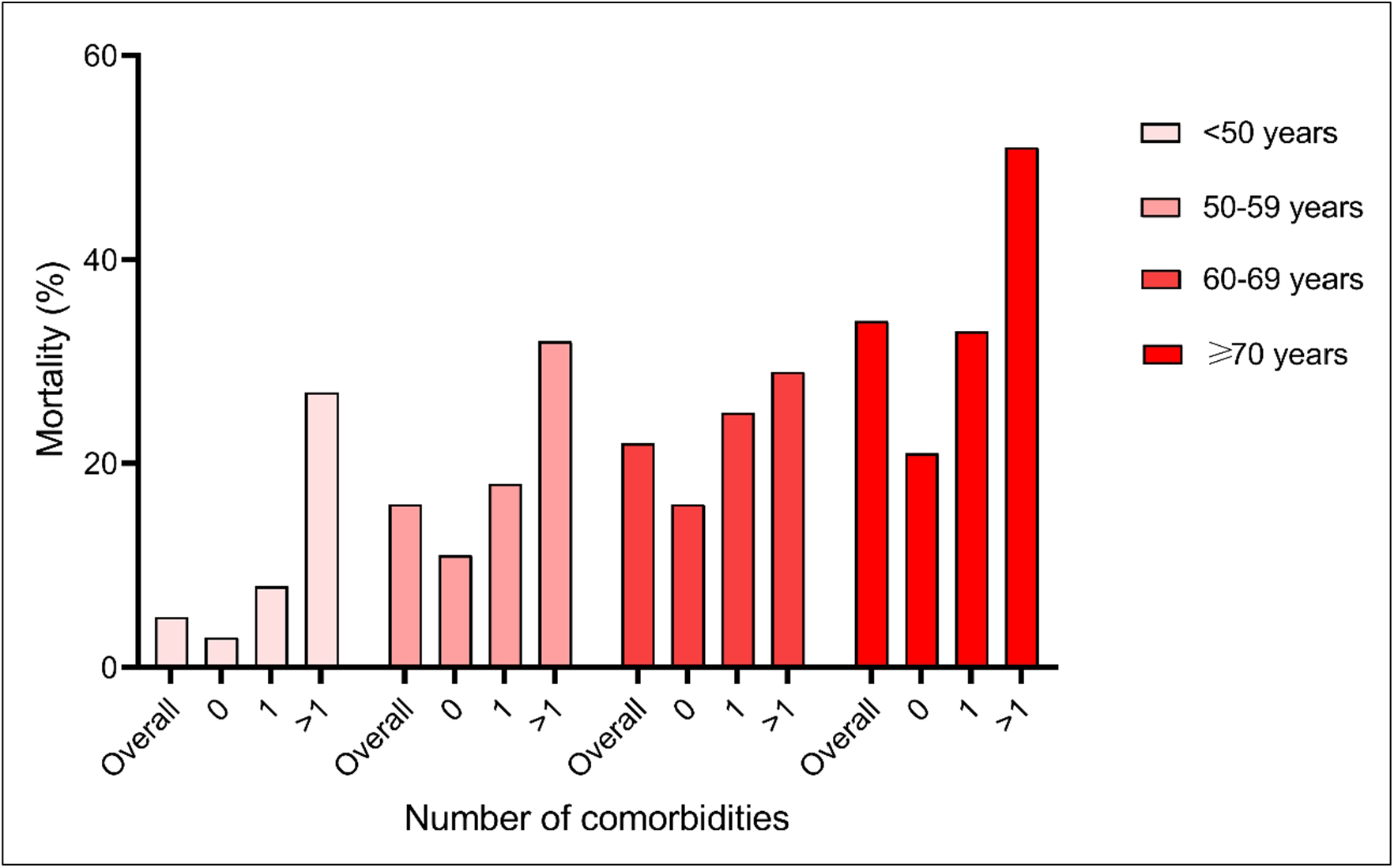
Age-specific mortality based on number of pre-existing comorbidities.

**Table S1:** Age-specific in-hospital mortality with confidence limits among patients hospitalised with COVID-19 in Jakarta, Indonesia.

|  | N total | N deceased | % deceased | 95% CI |
| --- | --- | --- | --- | --- |
| Age group (years) |  |  |  |  |
| 0-4 | 61 | 7 | 11.5 | 5.6-22.2 |
| 5-9 | 23 | 1 | 4.3 | 0.6-25.2 |
| 10-19 | 133 | 3 | 2.3 | 0.7-6.8 |
| 20-29 | 638 | 8 | 2.5 | 1.5-4.1 |
| 30-39 | 755 | 26 | 3.4 | 2.4-5.0 |
| 40-49 | 819 | 61 | 7.4 | 5.8-9.5 |
| 50-59 | 941 | 155 | 16.5 | 14.2-19.0 |
| 60-69 | 611 | 132 | 21.6 | 18.5-25.0 |
| 70-79 | 284 | 96 | 33.8 | 28.5-39.5 |

**Table S2:** Univariable analysis of demographic and clinical factors of mortality among patients hospitalised with COVID-19 in Jakarta, Indonesia.

|  | **Crude Hazard Ratio**<br>**(95% CI)** | **p value** |
| --- | --- | --- |
| Age group, years |  |  |
| <50 | 1 (ref) | .. |
| 50-59 | 4.61 (3.31-6.44) | <0.0001 |
| 60-69 | 5.14 (3.60-7.35) | <0.0001 |
| ≥70 | 7.72 (5.17-11.53) | <0.0001 |
| Sex |  |  |
| Female | 1 (ref) | .. |
| Male | 1.33 (1.04-1.70) | 0.024 |
| Number of symptoms |  |  |
| ≤3 | 1 (ref) | .. |
| >3 | 2.86 (2.16-3.77) | <0.0001 |
| Number of comorbidities |  |  |
| 0 | 1 (ref) | .. |
| 1 | 2.68 (1.98-3.62) | <0.0001 |
| >1 | 5.70 (4.25-7.66) | <0.0001 |
| Type of comorbidity |  |  |
| None | 1(ref) |  |
| Hypertension | 3.04 (2.37-3.91) | <0.0001 |
| Diabetes | 2.93 (2.22-3.85) | <0.0001 |
| Cardiac disease | 3.94 (2.92-5.31) | <0.0001 |
| COPD | 1.42 (0.84-2.40) | 0.186 |
| Chronic kidney disease | 5.32 (3.63-7.80) | <0.0001 |
| Immunocompromised | 3.05 (0.98-9.53) | 0.055 |
| Liver disease | 4.79 (1.78-12.89) | 0.002 |
| Malignancy | 1.03 (0.14-7.36) | 0.975 |
| Clinical diagnosis of pneumonia |  |  |
| Yes | 5.71 (4.21-7.73) | <0.0001 |
| No | 1 (ref) | .. |
| Immediate ICU admission |  |  |
| Yes | 7.23 (4.82-11.86) | <0.0001 |
| No | 1 (ref) | .. |
| Immediate intubation |  |  |
| Yes | 12.40 (8.19-18.77) | <0.0001 |
| No | 1 (ref) | .. |
| Time from symptom onset to admission, days |  |  |
| ≤7 | 1.58 (1.23-2.04) | <0.0001 |
| >7 | 1 (ref) | .. |
| Period of admission |  |  |
| March to April | 1.72 (1.31-2.27) | <0.0001 |
| May to July | 1 (ref) | .. |

## Notes

### Competing Interest Statement

The authors have declared no competing interest.

### Clinical Trial

NA

